# Enhanced differentiation of IgA^+^ class-switched CD27^-^CD21^+^ B cells in patients with IgA nephropathy

**DOI:** 10.1101/2024.04.29.24306572

**Authors:** Anna Popova, Baiba Slisere, Karlis Racenis, Viktorija Kuzema, Roberts Karklins, Mikus Saulite, Janis Seilis, Anna Jana Saulite, Aiga Vasilvolfa, Kristine Vaivode, Dace Pjanova, Juta Kroica, Harijs Cernevskis, Aivars Lejnieks, Aivars Petersons, Kristine Oleinika

**Affiliations:** Department of Nephrology, Pauls Stradins Clinical University Hospital, Riga, Latvia; Department of Biology and Microbiology, Riga Stradins University, Riga, Latvia; Department of Internal Medicine, University of Latvia, Riga, Latvia; Joint Laboratory, Pauls Stradins Clinical University Hospital, Riga, Latvia; Department of Doctoral Studies, Riga Stradins University, Riga, Latvia; Department of Internal Diseases, Riga Stradins University, Riga, Latvia; Institute of Microbiology and Virology, Riga Stradins University, Riga, Latvia; Riga East Clinical University Hospital, Riga, Latvia; Program in Cellular and Molecular Medicine, Boston Children’s Hospital, Harvard Medical School, Boston, MA, USA

**Keywords:** B cells, IgA nephropathy, IgA-producing plasmablasts, antibodies, disease mechanisms

## Abstract

**Background:** IgA nephropathy (IgAN) is characterised by the production of galactose-deficient IgA1 (Gd-IgA1) antibodies. As the source of pathogenic antibodies, B cells are central to IgAN pathogenesis, but the B cell activation pathways as well as the potential B cell source of dysregulated IgA-secretion remain unknown.

**Methods:** We carried out flow cytometry analysis of peripheral blood B cells in patients with IgA nephropathy and control subjects with a focus on IgA-expressing B cells to uncover the pathways of B cell activation in IgAN and how these could give rise to pathogenic GdIgA1 antibodies.

**Results:** In addition to global changes in the B cell landscape – expansion of naive and reduction in memory B cells – IgAN patients present with an increased frequency of IgA-expressing B cells that lack the classical memory marker CD27, but are CD21^pos^. IgAN patients further have an expanded population of IgA^pos^ antibody-secreting cells, which correlate with serum IgA levels. Both IgA^pos^ plasmabalsts and CD27^neg^ B cells co-express GdIgA1. Implicating dysregulation at mucosal surfaces as the driver of such B cell differentiation, we found a correlation between lipopolysaccharide (LPS) in the serum and IgA^pos^CD27^neg^ B cell frequency.

**Conclusion:** We propose that dysregulated immunity in the mucosa may drive de novo B cell activation within germinal centres, giving rise to IgA^pos^CD27^neg^ B cells and subsequently IgA-producing plasmablasts. These data integrate B cells into the paradigm of IgAN pathogenesis and allow to further investigate this pathway to uncover biomarkers and develop therapeutic interventions.

**Key learning points:** *What was known:*

- Patients with IgA nephropathy (IgAN) have aberrant production of galactose-deficient IgA1 (Gd-IgA1) and antibodies against it, which together form immune complexes that are deposited in the renal mesangium and lead to kidney damage; this is known as the multi-hit model of IgAN pathogenesis.
- The multi-hit model centrally implicates B cells as they produce both Gd-IgA1 and antibodies against it, yet B cell activation pathways that lead to aberrant antibody production are absent from the model.
- Only isolated reports exist describing specific features of B cells that are altered in patients with IgAN, including a reduction in regulatory B cells, increase in toll-like receptor 7 expression in total peripheral blood B cells and elevated frequency of circulating CCR9^+^IgA^+^ B cells.

*This study adds:*

- In addition to changes in the overall circulating B cell landscape, differentiation of IgA^+^ plasmablasts is enhanced in patients with IgAN and their levels correlate with serum IgA.
- IgA-expressing plasmablast frequency correlates with that of IgA^+^CD21^+^ B cells, that lack the classical memory B cell marker CD27.
- Both IgA^+^ plasmablasts and IgA-expressing CD27^-^ B cells co-express GdIgA1 receptors.
- IgA^+^CD27^-^CD21^+^ B cell frequency correlates with serum lipopolysaccharide (LPS) levels, implicating mucosa in their activation.

*Potential impact:*

- We uncover the previously unknown B cell activation pathway that appears to be associated with pathogenic IgA secretion in IgAN and integrate this into the multi-hit model of IgAN pathogenesis.
- This pathway holds potential for further investigation to identify biomarkers and therapeutic targets in IgAN.

## Introduction

IgA nephropathy (IgAN) is an autoimmune disease and the most common form of primary glomerulonephritis with an estimated global incidence of at least 2.5 cases per 100’000 adults annually (1). Patients present with heterogeneous clinicopathological manifestations and variable prognosis, from asymptomatic changes in urinalysis to rapidly progressive glomerulonephritis (2). Up to 39% of patients progress to end-stage renal disease (ESRD) over 20 years of follow-up (3). Present treatments carry severe side-effects and largely fail to halt the progression of renal decline (4). The mainstay of therapy is optimized supportive care, i.e., measures that lower blood pressure, reduce proteinuria, minimize lifestyle risk factors, and otherwise help to reduce non-specific insults to the kidneys. The use of immunosuppression has become controversial because of its low effectiveness and high rate of serious side effects mainly associated with glucocorticoid treatment (5). However new treatment regimens with low dose steroids and enteral targeted-release budesonide for high-risk patients may decrease the rate of IgAN progression (6). Treatment advances in IgAN have been limited at least in part due to the incomplete understanding of disease aetiopathogenesis.

The multi-hit model of IgAN aetiopathogenesis proposes that IgAN is initiated with the overproduction of galactose-deficient IgA1 (Gd-IgA1), followed by the development of antibodies against pIgA1 (7–10). Together these form immune complexes that lead to nephrotoxicity through complement activation via alternative and lectin pathways. B cells are centrally implicated in the pathogenesis of IgAN as the source of hypoglycosylated IgA1 and autoantibodies against it (11, 12). Nevertheless, there are crucial gaps in the knowledge on B cell activation and differentiation pathways that lead to secretion of pathogenic IgA antibodies in IgAN. This is particularly striking when compared to the comprehensive understanding gained in other immune-mediated kidney diseases, particularly systemic lupus erythematosus (13–15).

Isolated reports have characterised distinct aspects of B cell activation in patients with IgAN. Among the reported B cell alterations have been the reduction in regulatory B cells (16), elevated toll-like receptor 7 (TLR7) expression in circulating B cells (17) and increased frequency of CCR9^pos^IgA^pos^ B cells in blood (18). However, B cell activation pathways have not been investigated and remain to be integrated into the paradigm of IgAN aetiopathogenesis. Here we carry out peripheral B cell profiling in a biopsy-confirmed cohort of IgAN patients and healthy controls (HC) to uncover B cell activation pathways that operate in IgAN as well as to determine the cellular pathways implicated in the secretion of pathogenic IgA.

## Materials and Methods

### Study participants

Adults with biopsy-confirmed IgAN were recruited at the Nephrology Centre at Pauls Stradins Clinical University Hospital, Riga, Latvia between January 2020 and June 2022. This is the only centre for kidney diseases in Latvia, where IgAN is diagnosed in adults, therefore the IgAN patient cohort is representative of all Latvian adults with IgAN. Age- and sex-matched healthy volunteers were also enrolled. Healthy controls had age-appropriate kidney function, without active urine sediment and proteinuria. Individuals with diabetes mellitus, current pregnancy, severe organ dysfunction, acute cardiovascular disease, hepatic diseases, acute or chronic inflammatory, autoimmune or infectious diseases, immunodeficiency, malignancies, substance and alcohol abuse, kidney replacement therapy (including kidney transplantation) were excluded from the study. All participants provided written informed consent. This study was approved by the Clinical Research Ethics Committee of Pauls Stradins Clinical University Hospital (No 191219-6L) and was performed under the guidance of the Declaration of Helsinki.

### Clinical and laboratory characterisation

Serum creatinine, albumin and total cholesterol were measured on Atellica CH (Siemens Healthineers, Erlangen, Germany). Estimated glomerular filtration rate (eGFR) was calculated using the CKD-EPI Creatinine Equation (2021). Serum IgA was measured on Atellica NEPH 630 (Siemens Healthineers, Erlangen, Germany). Proteinuria was determined by spot protein-to-creatinine ratio. Assessment of protein in urine was performed on Cobas Integra 400 Plus (Roche Diagnostics GmbH, Mannheim, Germany). Red blood cell count in urine was determined on Atellica 1500 automated urinalysis system (Siemens Healthineers, Erlangen, Germany). The complete blood counts were performed on ethylenediaminetetraacetic acid (EDTA)-treated peripheral blood samples using UniCel DxH cellular analysis system (Beckman Coulter, Miami, FL, USA). Serum lipopolysaccharide (LPS) levels were detected by ELISA (MyBioSource MBS702450, San Diego, CA, USA) according to the manufacturer’s instructions. Gd-IgA1 levels in serum were measured using an ELISA kit (Gd-IgA1 Assay Kit-IBL 30111694, IBL international GmBH, Germany) following the manufacturer’s instructions. The samples were diluted 200-fold using the provided EIA buffer to obtain biomarker levels within the measurement range of the kit (1.56-100 ng/mL). Blood pressure was measured by a physician during study recruitment.

### Peripheral blood mononuclear cell (PBMC) isolation and serum collection

Peripheral blood was obtained from study participants. Serum from tubes with coagulation activator was isolated by centrifugation and was either used immediately or stored at -80C. PBMCs were isolated from heparinized blood by density gradient centrifugation using Histopaque-1077 (Sigma-Aldrich, St. Louis, USA). After washing with complete RPMI-1640 (10% fetal bovine serum and 1% penicillin-streptomycin in RPMI-1640), PBMCs were resuspended in freezing media (90% fetal bovine serum and 10% dimethyl sulfoxide) and cryopreserved. All laboratory analyses were done in the Joint Laboratory at Pauls Stradins Clinical University Hospital (Riga, Latvia).

### Immunophenotyping of peripheral blood B cells by flow cytometry

Using antibodies against CD24, CD27, CD38 and IgD we were able to enumerate transitional (CD24^hi^CD38^hi^), mature naive (CD24^int^CD38^int^), activated (CD24^lo^CD38^lo^), and total memory (CD24^hi^CD38^lo^) B cells, including class-switched (IgD^neg^) and unswitched (IgD^pos^) subsets, double negative (IgD^neg^CD27^neg^) B cells, pre-plasmablasts (CD24^lo^CD38^hi^) and plasmablasts (CD27^pos^CD38^hi^). We further interrogated IgA, CD21, T-bet, Ki-67 expression in B cell subsets. Briefly, for PBMC viability LIVE/DEAD Fixable Near-IR Dead Cell Stain Kit was used (Invitrogen, MA, USA). Nonspecific staining was prevented with Fc receptor blocking reagent (Miltenyi Biotec, Bergisch Gladbach, Germany). The cells were incubated with antibodies (details see Suppl. Table 1) for 40 minutes at 4C, afterwards unbound antibodies were removed by two washes with flow cytometry staining buffer (2% fetal bovine serum and 2 mM EDTA in phosphate-buffered solution (PBS)) and fixed with PBS containing 2% formaldehyde. For intracellular and transcription factor staining, cells were fixed and permeabilized using the Foxp3/Transcription Factor Staining Buffer Set (00-5523-00, eBioscience) followed by incubation with antibodies diluted in permeabilization buffer for 50 minutes at 4C. Samples were acquired on the Navios EX flow cytometer (Beckman Coulter, Inc., Brea, CA, USA) and analysed with FlowJo software (BD Life Sciences). For the detection of GdIgA1 B cells, we stained PBMCs with a fluorophore labelled (R10712 ReadyLabel™, Invitrogen, MA, USA) monoclonal antibody (Gd-IgA1 (KM55)-IBL 30117066, IBL Japan, Japan) at 37C for 40 minutes. To further enumerate Gd-IgA1 plasmablasts, cells were fixed again permeabilized using the Foxp3/Transcription Factor Staining Buffer Set and then stained intracellularly with the Gd-IgA1 monoclonal antibody.

### Statistical analysis

All the statistical analyses were conducted using GraphPad Prism 9 (La Jolla, CA, USA). Data distribution was assessed by the Shapiro-Wilk test and normal Q-Q plots. For normally distributed and homogenous data, independent samples t-test was used; when data were not normally distributed, Mann-Whitney U test was used. Fisher’s exact test was used to compare sex between groups. Spearman’s rank correlation test was used to interrogate statistical significance in correlations. Results were considered statistically significant at p<0.05.

## Results

To uncover B activation pathways and how peripheral B cell composition may be impacted in patients with IgAN, we recruited 36 patients with IgAN and 19 healthy controls (HCs). See Methods for full participant inclusion and exclusion criteria. The demographic, clinical and laboratory data of the cohort are summarised in Table 1. Study participants were sex- and age-matched. Both groups had normal and comparable leukocyte and lymphocyte counts. As expected, IgAN patients had higher serum creatinine levels and lower eGFR than HCs. IgAN patients represented all four CKD stages (from 1 to 4) based on eGFR. The median proteinuria of patients with IgAN was 0,48 g/g (IQR 0,26-1,35), 11 patients had moderate proteinuria (1-3g) and only one patient had nephrotic range proteinuria (>3g). According to the Oxford classification of IgAN (19), a frequent histological finding was secondary glomerulosclerosis (69,4%). Only in rare cases tubular atrophy or crescents in <25% of glomeruli were seen. Of note, BMI is associated with worse presentation and long-term outcome of IgAN (20, 21), and B cell activation pathways are dysregulated in obesity (22). In our cohort there were no significant differences in BMI between IgAN patients and controls.

**Table 1.**
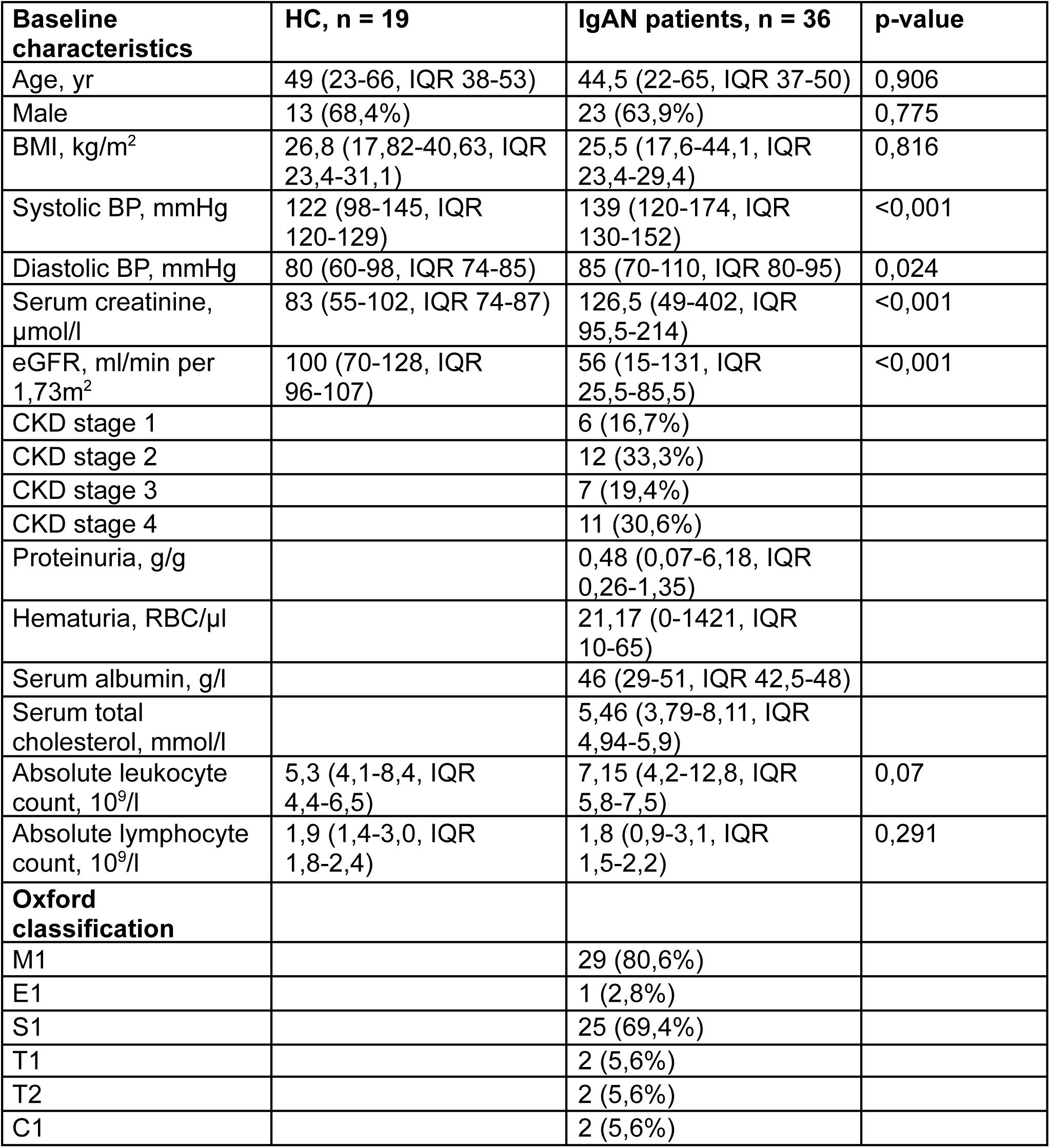
Demographic, clinical and laboratory characteristics of the cohort.

| **Baseline characteristics** | **HC, n = 19** | **IgAN patients, n = 36** | **p-value** |
| --- | --- | --- | --- |
| Age, yr | 49 (23-66, IQR 38-53) | 44,5 (22-65, IQR 37-50) | 0,906 |
| Male | 13 (68,4%) | 23 (63,9%) | 0,775 |
| BMI, kg/m<sup>2</sup> | 26,8 (17,82-40,63, IQR 23,4-31,1) | 25,5 (17,6-44,1, IQR 23,4-29,4) | 0,816 |
| Systolic BP, mmHg | 122 (98-145, IQR 120-129) | 139 (120-174, IQR 130-152) | <0,001 |
| Diastolic BP, mmHg | 80 (60-98, IQR 74-85) | 85 (70-110, IQR 80-95) | 0,024 |
| Serum creatinine, μmol/l | 83 (55-102, IQR 74-87) | 126,5 (49-402, IQR 95,5-214) | <0,001 |
| eGFR, ml/min per 1,73m<sup>2</sup> | 100 (70-128, IQR 96-107) | 56 (15-131, IQR 25,5-85,5) | <0,001 |
| CKD stage 1 |  | 6 (16,7%) |  |
| CKD stage 2 |  | 12 (33,3%) |  |
| CKD stage 3 |  | 7 (19,4%) |  |
| CKD stage 4 |  | 11 (30,6%) |  |
| Proteinuria, g/g |  | 0,48 (0,07-6,18, IQR 0,26-1,35) |  |
| Hematuria, RBC/μl | | 21,17 (0-1421, IQR 10-65) | |
| Serum albumin, g/l |  | 46 (29-51, IQR 42,5-48) |  |
| Serum total cholesterol, mmol/l |  | 5,46 (3,79-8,11, IQR 4,94-5,9) |  |
| Absolute leukocyte count, 10<sup>9</sup>/l | 5,3 (4,1-8,4, IQR 4,4-6,5) | 7,15 (4,2-12,8, IQR 5,8-7,5) | 0,07 |
| Absolute lymphocyte count, 10<sup>9</sup>/l | 1,9 (1,4-3,0, IQR 1,8-2,4) | 1,8 (0,9-3,1, IQR 1,5-2,2) | 0,291 |
| **Oxford classification** |  |  |  |
| M1 |  | 29 (80,6%) |  |
| E1 |  | 1 (2,8%) |  |
| S1 |  | 25 (69,4%) |  |
| T1 |  | 2 (5,6%) |  |
| T2 |  | 2 (5,6%) |  |
| C1 |  | 2 (5,6%) |  |

We first carried out B cell phenotyping based on CD24, CD27, CD38 and IgD surface expression. This allowed us to enumerate transitional (CD24^hi^CD38^hi^), mature naive (CD24^int^CD38^int^), activated (CD24^lo^CD38^lo^), and total memory (CD24^hi^CD38^lo^) B cells, including class-switched (IgD^neg^) and unswitched (IgD^pos^) subsets, double negative (IgD^neg^CD27^neg^) B cells, pre-plasmablasts (CD24^lo^CD38^hi^) and plasmablasts (CD27posCD38hi). The CD24/CD38 gating strategy confirmation is shown in Suppl. Figure 1. We found that IgAN patients had a significant increase in mature naive B cells with a reciprocal decrease in the frequency of total memory B cells (Figure 1a). Frequencies of switched and unswitched memory B cells were comparable (Figure 1b). A novel population of IgD^neg^CD27^neg^ B cells termed double negative (DN) or atypical memory B cells has been recently described. These DN B cells are expanded in autoimmune conditions, such as systemic lupus erythematosus (SLE), and especially in those with nephritis (14, 15). These cells have been shown to be the precursors of autoantibody-producing plasmablasts in SLE. Nevertheless, total double negative (DN) B cells and plasmablasts were comparable in patients with IgAN and HCs (Figure 1c).

**Figure 1.**
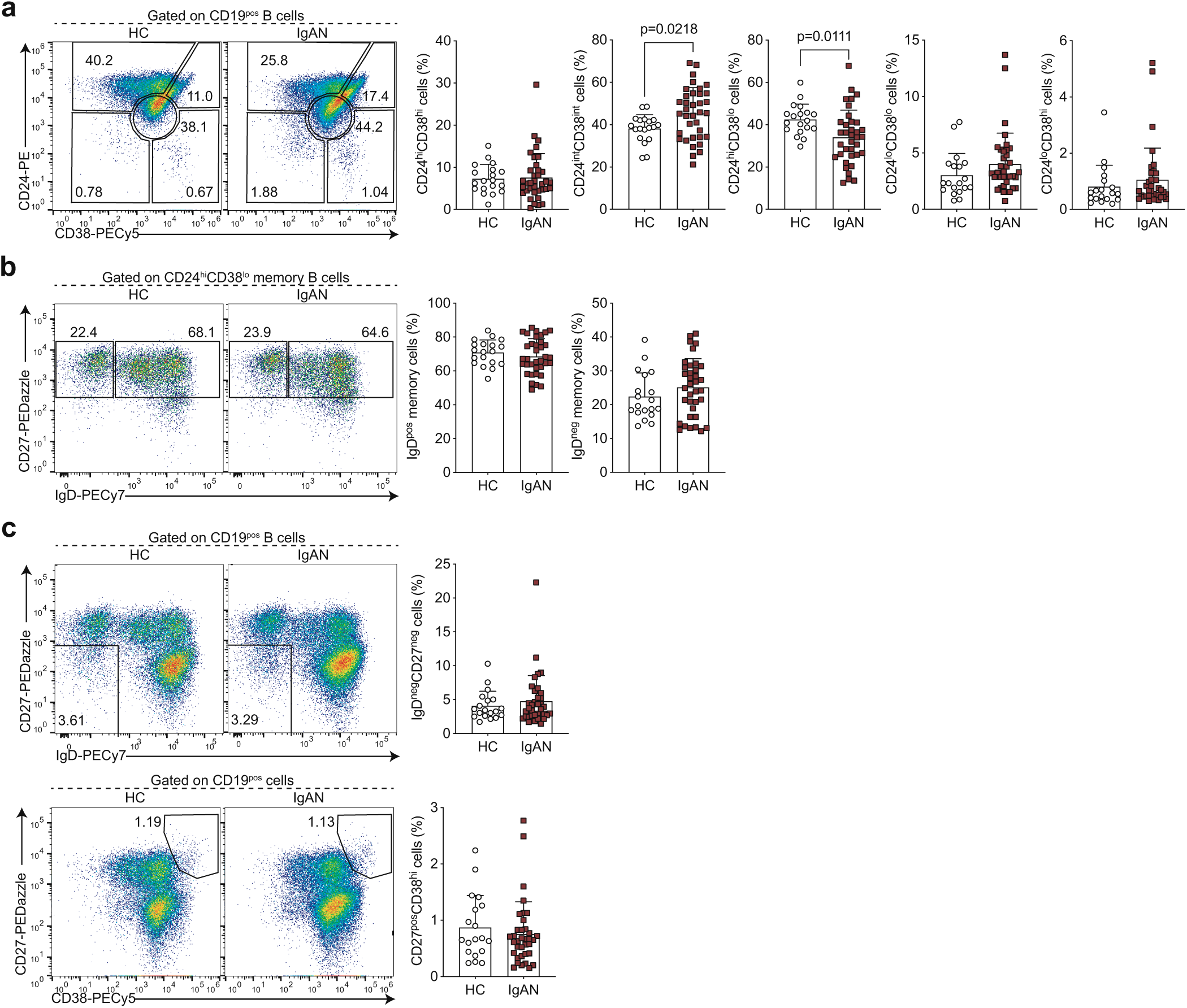
IgAN-associated changes in the peripheral B cell landscape. Representative flow cytometry plots and summary bar charts demonstrating (a) the frequencies of transitional (CD24^hi^CD38^hi^), mature (CD24^int^CD38^int^), memory (CD24^hi^CD38^lo^), and activated B cells (CD24^lo^CD38^lo^) and pre-plasmablasts (CD24^lo^CD38^hi^), (b) the distribution of memory B cells into IgD^pos^ unswitched and IgD^neg^ class-switched subsets, (c) the frequencies of total double negative (DN; IgD^neg^CD27^neg^) B cells and plasmablasts (CD27^pos^CD38^hi^) in IgAN patients and healthy controls. Data are mean±SD and each circle/square represents a study participant. For normally distributed CD24^int^CD38^int^, CD24^hi^CD38^lo^, IgD^pos^ memory and IgD^neg^ memory B cell populations independent samples t test was used to compare IgAN patients and healthy controls. For non-normally distributed CD24^hi^CD38^hi^, CD24^lo^CD38^lo^, CD24^lo^CD38^hi^, IgD^neg^CD27^neg^ and CD27^pos^CD38^hi^ B cell subsets Mann-Whitney U test was used for the comparison of IgAN patients and HCs.

The cellular origin and pathway that gives rise to IgA-producing B cells in IgAN is unknown, therefore, we next wanted to assay specifically IgA-expressing B cells and antibody-secreting cells (ASCs). That is, we wanted to know if systemic perturbations in the activation and differentiation of IgA-expressing B cells can be detected in patients with IgAN. Among B cells, IgA-expressing classical memory (CD27^pos^) B cell frequency was comparable between IgAN patients and HCs (Figure 2a). However, we noted that in addition to CD27-expressing B cells, there was a smaller population of IgA class-switched B cells that lacked CD27 expression, which was particularly pronounced in IgAN patients. Indeed, there was a significant expansion of these IgA-expressing CD27^neg^ B cells in patients with IgAN (Figure 2a). We also detected a significantly higher frequency of IgA class-switched ASCs in IgAN patients compared to controls (Figure 2b). Supporting a lineage relationship between IgA-expressing plasmablasts and IgA-expressing CD27^neg^ B cells, we found a correlation between these two subsets (Figure 2c).

**Figure 2.**
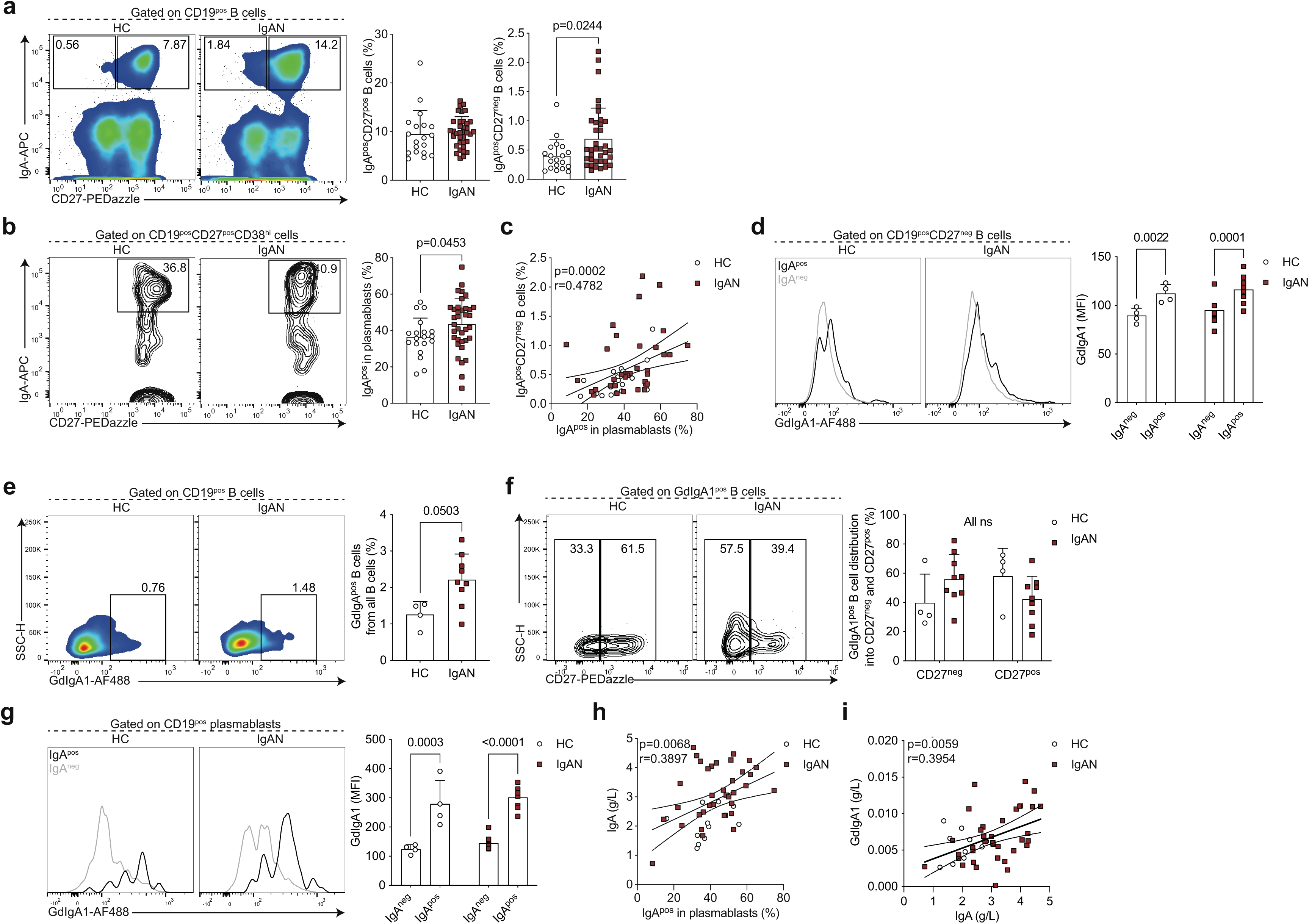
Enhanced differentiation of IgA^pos^CD27^neg^ B cells and IgA^pos^ plasmablasts in IgAN. Representative flow cytometry plots and summary bar charts demonstrating the frequencies of (a) IgA^pos^CD27^pos^ and IgA^pos^CD27^neg^ B cells and (b) IgA^pos^ plasmablasts, (c) linear regression analysis of IgA^pos^CD27^neg^ B cells versus IgA^pos^ plasmablasts, (d) representative flow cytometry histograms and summary bar charts demonstrating the median fluorescence intensity of GdIgA1 in IgA^pos^ and IgA^neg^ CD19^pos^CD27^neg^ B cells, (e, f) representative flow cytometry plots and summary bar charts demonstrating the frequencies of (e) GdIgA1^pos^ B cells and (f) the distribution of GdIgA1^pos^ B cells into CD27^neg^ and CD27^pos^ subsets, (g) representative flow cytometry histograms and summary bar charts demonstrating the median fluorescence intensity of GdIgA1 in IgA^pos^ and IgA^neg^ plasmablasts, (h, i) linear regression analysis of (h) serum IgA levels versus IgA^pos^ plasmablasts and (i) serum GdIgA1 versus serum IgA levels in IgAN patients and healthy controls. Data are mean±SD and each circle/square represents a study participant. For non-normally distributed subsets Mann-Whitney U test was used for the comparison of IgAN patients and HCs. Spearman’s rank correlation tests were used to interrogate statistical significance in the correlation between IgA^pos^CD27^neg^ B cells and IgA^pos^ plasmablasts, serum IgA levels and IgA^pos^ plasmablasts and serum GdIgA1 and serum IgA levels in IgAN patients and HCs.

We next wanted to address whether the IgA-expressing B cells were indeed expressing GdIgA1. We found that IgA^pos^CD27^neg^ B cells co-expressed GdIgA1 (Figure 2d). Reciprocally, in IgAN patients the majority of all GdIgA1^+^ B cells were CD27^neg^ (Figure 2e,f). IgA^pos^ plasmablasts also expressed high levels of GdIgA1 (Figure 2g). Finally, implicating the IgA-expressing plasmablast as a potential functional contributor to pathogenesis, we observed a correlation between IgA^pos^ plasmablasts and circulating IgA levels (Figure 2h). Nevertheless, despite the correlation between IgA and GdIgA1, we did not find a relationship between IgA^pos^ plasmablasts and serum GdIgA1 levels (Figure 2i).

CD27^neg^ antigen-experienced B cells comprise two subsets, termed double negative (DN) 1 and 2. DN1 cells are defined by their expression of CD21 and CXCR5, while DN2 B cells lack CD21 and CXCR5 expression and instead express CD11c and are transcriptionally regulated by T-bet (15). Based on their transcriptional signatures these two subsets of B cells arise from different activation pathways (15). While DN2 B cells arise through extrafollicular B cell activation, DN1 B cells represent precursors of classical memory B cells that have recently emerged from the germinal centre reaction (and have not yet upregulated CD27). Of note, germinal centres are the microanatomical structures that allow the evolution (by somatic hypermutation of antibody-encoding genes) and selection (affinity maturation) of B cells that enable the production of high-affinity antibodies (23). The understanding of which pathway B cells are activated through can elucidate factors that regulate the response (e.g. T cell help) or properties of the compartment (e.g. longevity) (23). We found that the IgA^neg^CD27^pos^ B cells were phenotypically CD21^hi^ and T-bet^lo^ corresponding to DN1 phenotype (Figure 3a), suggesting they may indeed be the precursors of IgA^pos^CD27^pos^ classical memory B cells. Of note, we used IgA^pos^CD27^pos^ memory B cells as a control for high expression of CD21 and lack of T-bet. To further interrogate this developmental relationship, we reasoned that most of the classical memory compartment (CD27^pos^) in patients with IgAN and HCs would be composed of foreign-antigen specific B cells generated throughout the lifetime. The vast majority of this pool should be resting cells, apart from those that report and participate in an on-going immune response. We then carried out Ki-67 staining and found a significant positive correlation between proliferating IgA^pos^ DN1 and Ki-67^pos^IgA^pos^CD27^pos^ memory B cells (Figure 3b). Therefore, the shared phenotypic and proliferation characteristics of these subsets suggest a developmental relationship and support increased generation of IgA-expressing ASCs through the germinal centre pathway.

**Figure 3.**
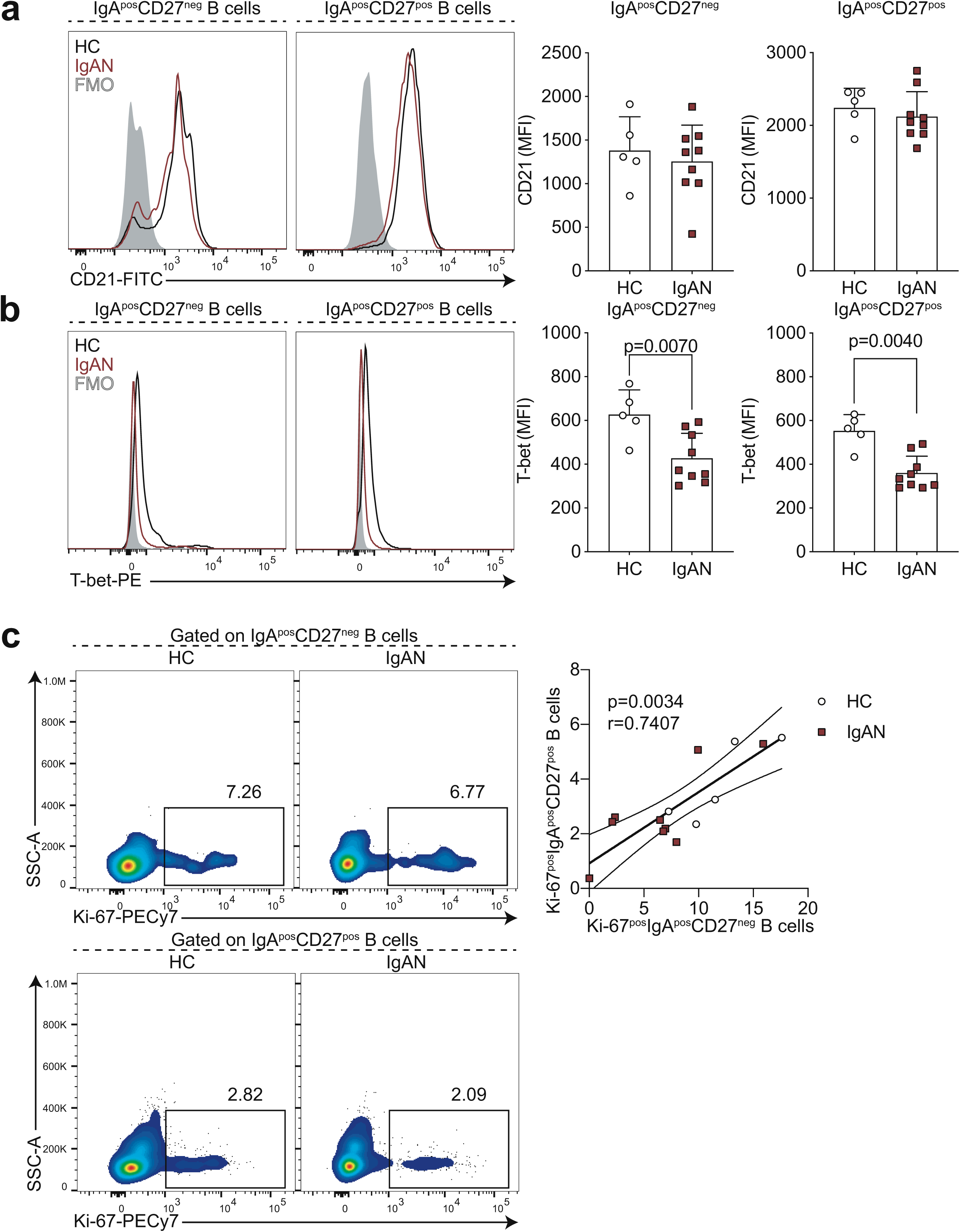
IgA^pos^CD27^neg^ B cells are phenotypically CD21^+^T-bet^-^. Representative flow cytometry histograms and summary bar charts demonstrating the median fluorescence intensity of CD21 (a) and T-bet (b) in IgA^pos^CD27^neg^ and IgA^pos^CD27^pos^ B cells, (c) representative flow cytometry plots and linear regression analysis of Ki-67^pos^IgA^pos^CD27^pos^ B cells versus Ki-67^pos^IgA^pos^CD27^neg^ B cells in HCs and IgAN patients. Data are mean±SD and each circle/square represents a study participant. For non-normally distributed CD21 and T-bet MFI Mann-Whitney U test was used for the comparison of IgAN patients and healthy controls. Spearman’s rank correlation test was used to interrogate statistical significance in correlations between Ki-67^pos^IgA^pos^CD27^pos^ B cells and Ki-67^pos^IgA^pos^CD27^neg^ B cells in IgAN patients and HCs.

Finally, we wanted to explore the mucosal-kidney axis in relation to B cell activation in IgAN and ask if previously reported perturbations at mucosal surfaces were linked to this DN1 B cell differentiation pathway. LPS is known to not only influence B cell class-switching to IgA (24, 25) but is also used as a surrogate marker of dysbiosis and gut permeability (26–29). We found that LPS was significantly elevated in the serum of IgAN patients compared to HCs (Figure 4a), confirming previously published data (30). We further found that IgA-expressing CD27^neg^ B cells correlated with serum LPS levels (Figure 4b; the relationships between LPS and the other B cell subsets examined in this study are presented in Suppl. Table 2). This suggests LPS may either directly drive their expansion/class-switching or that the observed correlation is because both increased LPS and IgA^pos^CD27^neg^ B cells are a consequence of mucosal dysbiosis/reduced barrier function, which contributes to IgA-expressing plasmablast differentiation. Finally, we wanted to ask how the expansion of IgA^pos^CD27^neg^ B cells related to clinical features of IgAN. We found that specifically in patients with reduced kidney function (eGFR <90mL/min) there was an inverse correlation between eGFR and the frequency of IgA^pos^CD27^neg^ B cells (Figure 4c).

**Figure 4.**
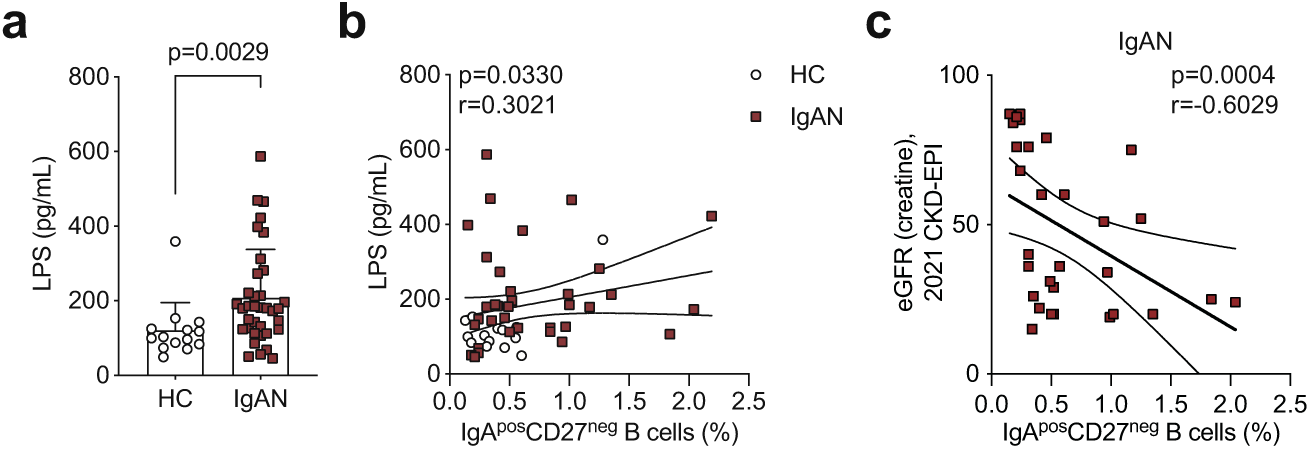
Serum LPS levels correlate with IgA^pos^CD27^neg^ B cell frequency. (a) Summary bar chart showing serum LPS levels and (b) linear regression analysis of serum LPS levels versus IgA^pos^CD27^neg^ B cells in IgAN patients and HCs. (c) Linear regression analysis of eGFR versus IgA^pos^CD27^neg^ B cells in IgAN patients. Data are mean±SD and each circle/square represents a study participant. For non-normally distributed serum LPS levels Mann-Whitney U test was used for the comparison of IgAN patients and healthy controls. Spearman’s rank correlation test was used to interrogate statistical significance in the correlation between serum LPS levels and IgA^pos^CD27^neg^ B cells in IgAN patients and HCs.

## Discussion

The multi-hit model is the blueprint for explaining IgAN aetiopathogenesis (7–10). Advances have been made particularly in uncovering the mechanisms operating within the kidney that contribute to organ damage (hit 4, see Figure 5). These include the dissection of how immune complexes containing IgA bind to mesangial cells, triggering proliferation and increased synthesis of extracellular matrix components as well as the role of the CD89, the IgA Fc receptor, in regulating tissue damage (31, 32). However, these mechanistic insights are unlikely to provide the key to therapeutic advances as they operate once B cell tolerance is already broken.

**Figure 5.**
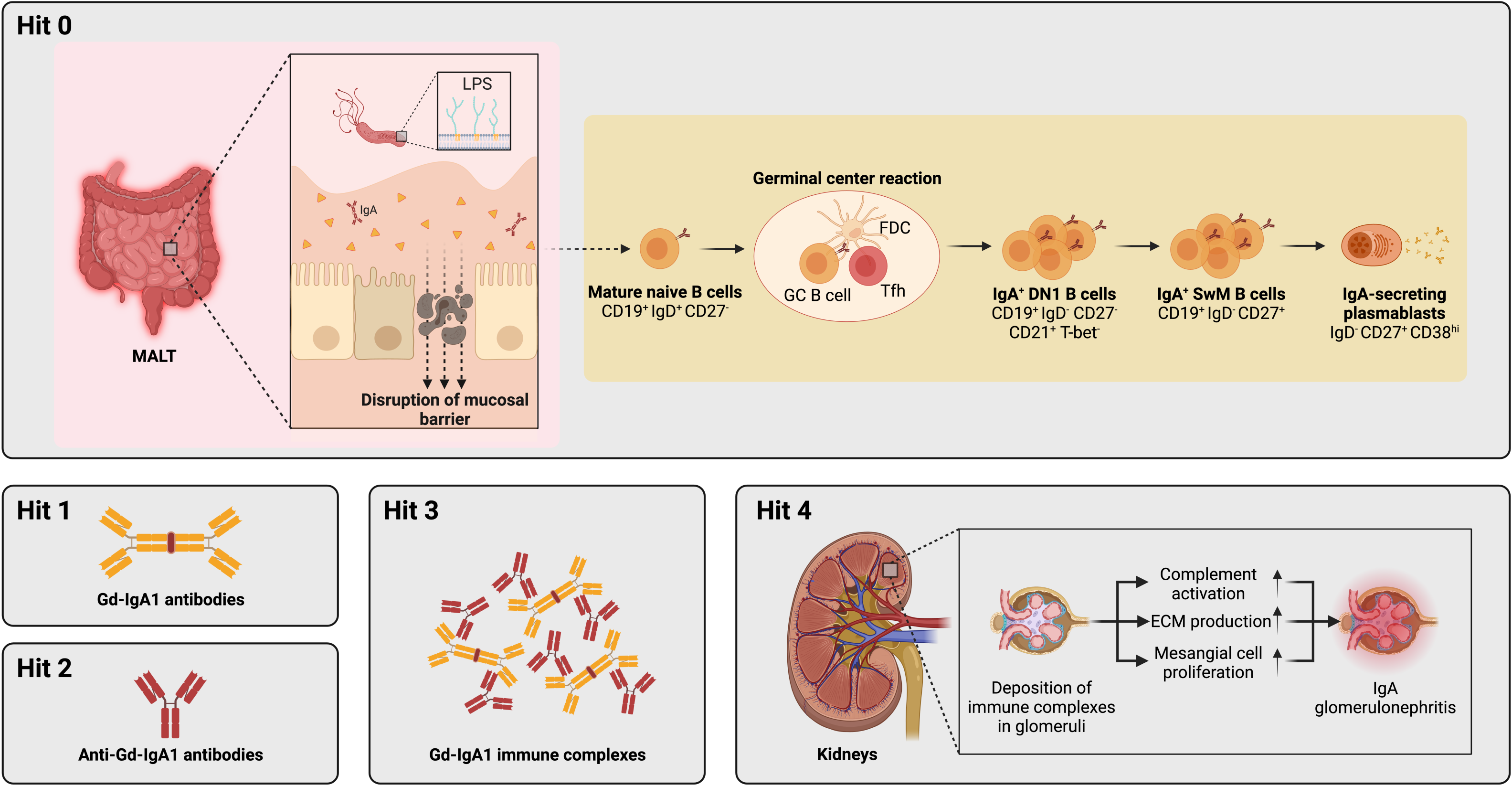
Revised multi-hit model with integration of B cell activation pathways. Hit 1 of the multi-hit pathogenesis is the production of pathogenic galactose-deficient IgA1 (Gd-IgA1). This is followed by production of anti-IgA antibodies (Hit 2) and formation of immune complexes (Hit 3), which are finally deposited in the kidney glomeruli and cause tissue damage and renal decline. Here we have uncovered the B cell pathway that leads to Gd-IgA1 production (hit 1) in IgAN pathogenesis, and term it ‘hit 0’. Specifically, naive B cells are de novo activated within germinal centers and give rise to IgA^pos^CD27^neg^ DN1 B cells and subsequently IgA-expressing plasmablasts which secrete pathogenic IgA.

We report here that global alterations in the B cell compartment are present in patients with IgAN, specifically the expansion of naive B cells and a reduction in total memory B cells. This observation is of significance as IgAN is considered an organ-specific autoimmune disease, yet these data support that systemic immunity is dysregulated beyond that which has been previously described at mucosal sites (33–35). Increase in naive B cells and reduction in total memory B cells is characteristic of systemic autoimmune diseases such as systemic lupus erythematosus (SLE) and conditions associated with chronic low-grade inflammation, including obesity (15, 22). However, we did not observe an expansion of total DN B cells associated with most severe systemic autoimmune and inflammatory phenotypes. It will be important to address whether the systemic changes in the overall B cell landscape we report here are causal to IgAN or a consequence of autoimmunity-associated inflammation/bystander effects.

As the source of pathogenic Gd-IgA1 antibodies, B cells are a promising target in IgAN, but B cell activation and differentiation pathways that lead to pathogenic IgA production remain uncharacterised. Understanding how B cells are affected and become activated is thus important for the complete understanding of IgAN pathogenesis, prognosis, and treatment. Here we provide mechanistic insight into the early steps of IgAN aetiopathogenesis by uncovering that IgA-expressing B cells that lack the classical memory marker CD27 but express CD21 are expanded in patients with IgAN and appear to be the cellular precursors of IgA-producing plasmablasts. We show that such ASC precursors are apparently recent emigrants from on-going germinal centres, which suggests the continuous output of the pathogenic precursors, rather than dominant contribution by memory B cells and plasmablasts differentiated at an earlier timepoint. DN1 B cells have not been previously associated with disease states (unlike the extensively characterised DN2 subset) therefore, this work may help uncover more about this B cell population. This observation of increased recent germinal centre emigrant DN1 B cells also fits with the report of increased frequency of Tfh cells in IgAN patients (36).

That we found a correlation between the CD27^neg^ precursors of IgA-expressing plasmablasts and serum LPS levels, implicates mucosal immunity in the differentiation of these cells. LPS could directly contribute to class-switching in activated B cells as previously observed (24, 25). In support of direct activation, tonsillar B cells from IgAN patients have been shown to produce increased IgA in response to LPS stimulation (37). Others have shown that upon in vitro culture with LPS, B cells downregulate the expression of β3-galactosyltransferase-specific molecular chaperone (Cosmc) (38). Therefore, LPS may act at multiple stages of B cell activation to not only induce class-switching and IgA production, but to also regulate IgA glycosylation. Another possible explanation for the correlation between IgA^pos^CD27^neg^ B cells and LPS is that both are increased as a consequence of intestinal inflammatory processes that may result in enhanced generation of IgA^pos^CD27^neg^ B cells and increased intestinal permeability. Supporting the mucosa as a site of origin for these IgA-expressing B cells, IgAN patients have an increased frequency of IgA^pos^ B cells that express the mucosal homing marker CCR9 (18). It is possible that tonsillectomy, a procedure that has demonstrated to be of some benefit in IgAN (39, 40), may eliminate the niche of either generation or homing of these B cells as has been speculated by others (41). It may also provide clues as to why B cell depletion has shown variable success in IgAN despite the central role of B cells in the disease pathogenesis (42)– the mucosal niches may harbour the majority of precursors that are not efficiently depleted by the B cell targeting therapy. Albeit the literature is not consistent, studies have suggested that rituximab may be less effective in depleting B cells within lymphoid tissues (43). Recent findings of disease-modifying effects of enteral budesonide in IgAN further implicate mucosal sites in driving immunopathogenesis (44). Whether budesonide affects B cell phenotypes remains to be determined, but it is tempting to speculate that it indirectly targets IgA^pos^ ASC precursors at mucosal sites. Additionally in other autoimmune diseases both memory B cells and B cells de novo engaging in immune responses – such as germinal centre B cells which are the proposed progenitors of DN1 B cells – have been shown to be resistant to rituximab treatment (45, 46).

In IgAN endocapillary proliferation, tubular atrophy/interstitial fibrosis are independent predictors of rate of loss of renal function (47). In our cohort of IgAN patients, the prevalence of these findings was relatively low. It is important to note that kidney biopsy may have been conducted several years before the patient’s visit. The distribution of men, as well as the age and BMI, was found to be similar to that reported in other studies focusing on IgAN. Clinically, patients exhibited lower protein excretion, yet their eGFR was worse when compared to the results of other studies evaluating B cells in IgAN (16, 18). Of note, a limitation of the study is that our cohort consisted entirely of white Eastern Europeans and thus the B cell activation pathway we have uncovered remains to investigated in non-European cohorts.

Now that we have identified the aberrant B cell differentiation pathways in IgAN, important further questions can be addressed. Due to their expression of GdIgA1, it is tempting to speculate that IgA^pos^CD27^neg^ cells participate in disease precipitation, albeit this remains to be experimentally demonstrated. That is, the enhanced generation of IgA^pos^CD27^neg^ precursors could drive disease or these cells may be generated as part of ongoing inflammation and not causal in IgAN development. Since this study aimed to capture the whole range of IgAN clinical features, our cohort comprised only 6 patients with eGFR>90mL/min. We speculate that if the B cell dysregulation we characterise here underlies disease, patients would present with an increased activation of this pathway prior to detectable decline in renal function. With regards to the antigen specificity and clonality of IgA-expressing B cells: what are the B cells recognising (e.g. food, microbial antigens) and how diverse is the IgA response within an individual and between individuals? With regards to the site of induction and long-term residence – where are the IgA-expressing plasmablast precursors induced and maintained (e.g. mucosa, bone marrow) and is this shared across IgAN patients? How are pathogenic IgA^pos^CD27^neg^ B cells different from foreign-antigen reactive IgA^pos^CD27^pos^ B cells– can targets be identified to specifically deplete or inhibit the differentiation of pathogenic B cells?

In summary, we propose that dysregulation of mucosal immunity may drive the increased naïve B cell activation in germinal centres, giving rise to IgA^pos^CD27^neg^CD21^pos^ B cells and subsequently IgA-producing plasmablasts; this pathway can be further explored for biomarkers and therapeutic targets in IgAN.

## Data availability statement

The data that support the findings of this study are available in the article and in its online supplementary material.

## Acknowledgements

We thank Dr Elizabeth C. Rosser for critically reviewing the manuscript and all the study participants, who made this research possible.

## Authors‘ contributions

AP, KR, VK, JK, AL, KO designed the study; AP, KR, VK, HC, AP were responsible for patient and control group selection and enrolment; AP, KR, VK, MS, JS, AJS, AV collected patient medical histories, obtained patient samples; AP, BS, KR, VK, RK, MS, JS, AJS, AV, KV, DP, JK, HC, AL, AP, KO conducted experiments and analysed data; KO and RK interpreted results, wrote and revised the manuscript and all authors contributed to the final manuscript.

