## Supplemental Figure 1 for "Enhanced differentiation of IgA^+^ class-switched CD27^-^CD21^+^ B cells in patients with IgA nephropathy"

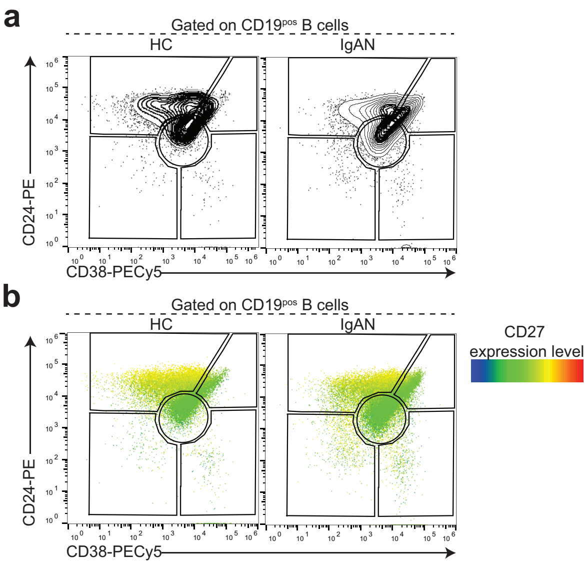


**Suppl. Figure 1.** CD24/CD38 gating strategy. Representative flow cytometry plots demonstrating the distribution of transitional (CD24$^{hi}$CD38$^{hi}$), mature (CD24$^{int}$CD38$^{int}$), memory (CD24$^{hi}$CD38$^{lo}$), and activated B cells (CD24$^{lo}$CD38$^{lo}$) and pre-plasmablasts (CD24$^{lo}$CD38$^{hi}$) as contour (a), and as CD27 heatmap statistic (b) plots in healthy controls and patients with IgAN.
