## Supplemental Table 1 for "Enhanced differentiation of IgA^+^ class-switched CD27^-^CD21^+^ B cells in patients with IgA nephropathy"

**Suppl. Table 1.** Antibodies used in flow cytometry.

| Antigen | Fluorochrome | Clone | Cat. no. | Supplier | Dilution |
|---|---|---|---|---|---|
| CD3 | APC-Cy7 | HIT3a | 300318 | BioLegend | 1:20 |
| CD19 | AF700 | HIB19 | 557921 | BD Biosciences | 1:50 |
| CD21 | FITC | Bu32 | 354910 | BioLegend | 1:20 |
| CD24 | PE | ML5 | 311106 | BioLegend | 1:50 |
| CD27 | PE-Dazzle 594 | M-T271 | 356422 | BioLegend | 1:50 |
| CD38 | PE-Cy5 | HIT2 | 303508 | BioLegend | 1:20 |
| IgA | APC | IS11-8E10 | 130-113-472 | Miltenyi Biotec | 1:50 |
| IgD | FITC | IA6-2 | 555778 | BD Biosciences | 1:50 |
| Ki-67 | PE-Cy7 | 20Raj1 | 25-5699-42 | eBioscience | 1:20 |
| T-bet | PE | 4B10 | 12-5825-82 | eBioscience | 1:20 |
