## Supplemental Table 2 for "Enhanced differentiation of IgA^+^ class-switched CD27^-^CD21^+^ B cells in patients with IgA nephropathy"

**Suppl. Table 2.** Relationships between serum LPS levels and B cell subsets.

CD27+CD38++IgA+CD27+: R=0.201, p=0.162

CD24loCD38lo: R=0.34, p=0.016*

IgA+CD27+: R=0.201; p=0.163

CD24intCD38int: R=0.185, p=0.291

CD24hiCD38lo: R=-0.213, p=0.137

CD24hiCD38lo/CD27+IgD+: R=0.258, p=0.07

CD24hiCD38lo/CD27+IgD-: R=-0.263, p=0.65

CD24hiCD38loIgA+: R=0.232, p=0.105

CD24hiCD38lo/IgA+CD27+: R=0.228; p=0.112

CD24hiCD38lo/IgA+CD27-: R=0.037, p=0.801

Mature CD24intCD38int: R=0.215; p=0.134

MemoryCD24hiCD38lo: R=-0.148; p=0.305

Transitional CD24hiCD38hi: R=-0.177; p=0.219

Transitional CD24hiCD38hi: R=-0.115, p=0.426
